# MEDIAL FRONTAL THETA (MFT) AS A PREDICTOR OF ANXIETY SENSITIVITY TREATMENT RESPONSE

**DOI:** 10.1101/2024.10.23.24313013

**Authors:** Jessica Ellis, Glenda Mello, Devin Butler, N. B. Schmidt, Edward Bernat

**Affiliations:** University of Maryland - College Park; Florida State University

## Abstract

Despite various interventions for anxiety disorders, effects can take months and many individuals do not respond. Activity in the anterior cingulate cortex is a consistent predictor of treatment outcomes, and can be measured by medial frontal theta (MFT) event-related potentials. This study used task-based electroencephalography and MFT to predict anxiety sensitivity treatment response at mid-treatment, 1-week post-treatment, and 6 months post-treatment. Results indicated that lower medial to lateral prefrontal theta phase synchrony was associated with greater symptom improvement. This work represents a novel finding that may contribute to the improvement in treatment efficacy by serving as a target for future interventions and individualized treatment selection.

## Introduction

### Anxiety and its impact on society

Anxiety is one of the most prevalent mental health problems in the United States and around the world (Collins et al., 2011; Kessler, Petukhova, Sampson, Zaslavsky, & Wittchen, 2012). According to data provided in the National Comorbidity Survey, four of the five most prevalent lifetime disorders are anxiety disorders: specific phobia (15.6%), social phobia (10.7%), post-traumatic stress disorder (PTSD; 5.7%) and generalized anxiety disorder (GAD; 4.3%).

With such high prevalence rates, the effects of anxiety on the individual and society remain substantial and can be very debilitating. For instance, it has been suggested that the level of social disability associated with generalized anxiety disorder (GAD) is as severe as seen with chronic somatic diseases (Kessler et al., 2001), and approximately 34% of patients with GAD will show a reduction of work productivity of 10% or more in a month (Witcchen et al., 2000). Economically, anxiety disorders contribute significantly to mental health costs with some estimates suggesting that anxiety disorders cost $46.6 billion in direct (i.e., patient visits, professional services, medication costs, etc.) and indirect costs (i.e., lost income, lost productivity, mortality etc.), accounting for 31.5% of the total cost of mental illness in America (DuPont et al., 1996). As such, there is a need for more research in this domain, particularly studies that may offer insight into anxiety treatment outcomes.

### Importance of predicting treatment response

Despite effective pharmacotherapy treatments for anxiety, response rates are estimated at approximately 60%, indicating that nearly half of the individuals undergoing treatment do not show any change in symptom severity (Nimatoudis et al., 2004; Pollack, 2001). Consequently, there is a critical need for more effective treatments, and more informed and individualized treatment selection (Simon & Perlis, 2010). Since many treatments take weeks or months to see effects, patients and clinicians would benefit from having objective pre-treatment measures that predict treatment response.

Several sociodemographic and clinical factors have been identified in this regard, including history of failed treatments (Ellis, Zarate, Luckenbaugh, & Furey, 2014), pre-treatment symptom severity (Connor, Hidalgo, Crockett, Malik, Katz, & Davidson, 2001; Doehrmann et al., 2013; Karatzias et al., 2007; Otto, Pollack, Gould, & Worthington, 2000; Tukel, Bozkurt, Polat, Genc, & Atli, 2006), comorbidity with other disorders (Baer, Jenike, Black, Treece, Rosenfeld, & Greist, 1992), and avoidant personality traits (Chambless, Tran, & Glass, 1997). Additionally, biological predictors of treatment response may offer greater insight into the neurobiological substrates of the disorders and serve as potential targets for future interventions.

### Anxiety Sensitivity

Anxiety sensitivity is the fear of anxiety-related symptoms (e.g sweating, increased heart rate) and it has been shown to be predictive of development and persistence of anxiety disorders (Deacon and Abramowitz, 2006; Hovenkamp-Hermelink et al., 2019, Reiss and McNally, 1985). Early work investigating AS demonstrated contributions to the etiology and maintenance of panic disorder (PD; Schmidt, Lerew, & Jackson, 1997; 1999). Recently, AS has also been implicated in the development of several other affective disorders (Schmidt, Zvolensky, & Maner, 2006), including depression and generalized anxiety disorder (Allan, Capron, Raines, & Schmidt, 2014), obsessive-compulsive disorder (OCD; Raines, Oglesby, Capron, & Schmidt, 2014), posttraumatic stress disorder (PTSD; Lang, Kennedy, & Stein, 2002), substance use disorders (McCaul, Hutton, Stephens, Xu, & Wand, 2017; Paulus, Hogan, & Zvolensky, 2018), and externalizing disorders (Bilgic et al., 2017). Further, empirical evidence suggests AS can be treated through brief interventions, ultimately resulting in reduced symptom severity across psychopathologies (Schmidt et al., 2007; Schmidt, Capron, Raines, & Allan, 2014; Smits et al., 2008).

Evidence also points to AS as an excellent target to examine treatment related changes. For instance, changes in AS have been shown to completely mediate change in some CBT trials of anxiety psychopathology (Smits et al., 2004; Smits et al., 2008). Additionally, a recent meta-analysis by Fitzgerald and colleagues (2021), which evaluated AS as a transdiagnostic risk factor and potential treatment target, concluded that AS can be approached as a modifiable mechanistic target. Various studies have also suggested that AS can be treated through brief interventions, ultimately resulting in reduced symptom severity across psychopathologies, and implicating AS as a reliable proxy to show overall symptomatic improvement. (Fitzgerald et. al, 2021; Schmidt et al., 2007; Schmidt, Capron, Raines, & Allan, 2014; Smits et al., 2008)

There has also been some evidence pointing to the neural processes specifically associated with AS severity. Results from these fMRI studies show a positive association between self-reported AS and activity in the insula and anterior cingulate cortex (ACC) during emotional processing tasks (Poletti et al., 2015; Stein et al., 2007).

### Neuroimaging predictors of treatment response: Depression

The majority of work identifying neuroimaging predictors of treatment response has been done in Major Depressive Disorder (MDD) and pre-treatment ACC activity has been shown to consistently predict treatment outcomes. This has been demonstrated by studies using PET, fMRI and EEG across various tasks and has been consistently found as a response to different antidepressant treatments (Jaworska et al. 2023; Keedwell et al., 2010; Pizzagalli et al. 2018; Saxena et al., 2003; Spronk et al. 2011) as well as to Cognitive Behavioral Therapy (CBT) (Feurer et al. 2022; McGrath et al., 2014; Siegle et al., 2006, 2012). Such findings may potentially be used to optimize treatment selection by differentiating between medication and psychotherapy response.

### Neuroimaging predictors of treatment response: Anxiety

Because depression is often comorbid with anxiety, we might expect to see similar results for predicting anxiety treatment response. Even though research in this area is nascent, emerging evidence shows a similar effect, indicating the ACC is the most consistent predictor for anxiety treatment outcomes as well (Ball, Stein, & Paulus, 2014; Shin et al., 2013).

Of the existing studies, converging evidence suggests greater dorsal ACC consistently predicts better treatment response (Hendler et al., 2003; Swedo et al., 1989). However, the mechanisms of this effect are still not clear (e.g., functional connectivity to DLPFC) and the direction of the effect may depend on the type of anxiety disorder (i.e., less activation for obsessive compulsive disorder (OCD), but greater activation for all other anxiety disorders). Moreover, a majority of previous work has been limited to fMRI or PET paradigms. While not surprising given the use of these methods during the inception of treatment response work, EEG may serve as a more cost effective method with potentially greater applicability in clinical settings. As such, more work is needed to clarify and expand upon the role of the ACC in anxiety treatment response.

### Anterior Cingulate Cortex (ACC) and Medial Frontal Theta

Midfrontal theta (MFT) power can be characterized as a marker of cognitive processing and cognitive control because of its association with cognitive conflict and error detection (Cavanagh & Shackman, 2015; Nigbur et al., 2011; McLoughlin et al. 2022). MFT activity has been widely observed to rely on the ACC to communicate with sensory and motor regions when responding to conflict or error (Cavanaugh and Frank, 2014; Cohen et al., 2008; Hauser et al., 2014; Sipp et al., 2013). A meta-analysis by Shackman and colleagues suggested that the ACC integrates negative affect, pain, and cognitive control, functioning as a hub that modulates behavior by interacting with other brain regions, potentially explaining the relationship between greater ACC activation and better treatment outcomes (Shackman et al., 2011). Activity in the ACC can be reliably measured by EEG and medial frontal ERPs, including the Feedback Negativity (FN), Error Related Negativity (ERN), and “control” N2, designated MFT negativities (Cavanagh et al., 2012; Cavanagh & Shackman, 2015; Hauser et al., 2014; Miltner, Braun, & Coles, 1997).

### Medial Frontal Theta (MFT) and anxiety

Medial frontal theta ERPs (i.e., ERN, FN, & N2) are consistently shown to reflect activity in ACC, and are generally enhanced in relation to various measures of anxiety (Banica et al., 2020; Cavanagh & Shackman, 2015; Ellis, Watts, Schmidt, & Bernat, 2018; Hajcak and Olven, 2008; Meyer et al. 2015; Moser et al. 2013; Mueller et al., 2015; Osinsky et al., 2017).

However, while extensive implicates ACC in prediction of treatment response, medial frontal theta ERPs have not been evaluated as predictors of treatment response. Consistent with the emerging interest in treatment response for anxiety, examining these components represents a novel and important contribution to this area. As such, our primary aim is to evaluate medial frontal theta ERPs as predictors of anxiety sensitivity treatment response.

### Medial Frontal Theta as a shared process

Given that all three medial frontal theta ERPs have been shown to reflect activity in ACC, and all three have demonstrated a similar association with anxiety, it has been suggested that they may represent a shared process in relation to motivationally significant outcomes (Cavanagh et al., 2012; Cavanagh & Shackman, 2015,Holroyd et al., 2008, Holroyd & Coles, 2002). A number of studies support the view that all three components reflect activity in medial frontal regions (Cavanagh & Shackman, 2015; Gehring & Willoughby, 2002; Hauser et al., 2014; Holroyd et al., 2004; Miltner, Braun, & Coles, 1997; Potts, Martin, Burton, & Montague, 2006). However, while the ERN and FN are both thought to reflect a performance monitoring system with primary activity over medial frontal regions, evidence for at least partially distinct processes underlying these components has been shown in terms of differing scalp distributions (Foti, Weinberg, Bernat, & Proudfit, 2014; Gehring & Willoughby, 2004) and differential sensitivity to externalizing psychopathology (Bernat, Nelson, Steele, Gehring, & Patrick, 2011), obsessive-compulsive traits (Simons, 2010), and schizophrenia (Horan, Foti, Hajcak, Wynn, & Green, 2012).

Notably, there has been a recent increase in research investigating the associations between MFT and psychopathology. Because MFT plays a central role in many cognitive control deficits associated with various psychiatric disorders, it has been suggested that MFT, indexed by ERPs, can be essential to understanding the underlying processes associated with psychopathology vulnerabilities (Buzsák and Watson, 2012; McLoughlin et al., 2022). The ERN specifically is being proposed as a potential transdiagnostic marker for various psychological disorders (McLoughlin et al., 2022; Passion and Barbosa, 2019). Research consistently shows a smaller ERN amplitude in relation to externalizing disorders and larger amplitude in relation to internalizing disorders (Banica et al., 2020; Hajcak and Olven, 2008; Pasion and Barbosa, 2019; Matthews et al., 2012; Moran et al., 2017; Moser 2016; Riesel 2019). However, it is important to note that findings extend to overall midfrontal theta, identifying additional theta-related ERPs as potential markers of anxiety, OCD, ADHD and substance use (Cavanagh and Shackman 2015; Cavanagh et al., 2017; Geburek et al., 2013; Harper et al., 2021; Luijten et al. 2014; Riesel et al., 2017; Rommel et al., 2019; Schmidt et al., 2018; Riesel et al., 2017). These findings have also broadened beyond the internalizing and externalizing spectrum, with a decrease in midfrontal theta activity being associated with schizophrenia (Ryman et al., 2018; Boudewyn and Carter, 2018). Thus, MFT has been broadly implicated in psychopathology, with some strong shared relationships across MFT generating tasks. This work supports the view that MFT is broadly decreased with regard to externalizing and psychotic disorders, and increased relative to anxiety.

### Medial Frontal Theta: Medial-Lateral Functional Connectivity

To index functional connectivity between medial and lateral frontal areas, we employed time-frequency based inter-channel phase synchrony (ICPS) (Aviyente et al., 2011). Work from our group and others have demonstrated sensitivity of medial-lateral functional connectivity measures to control functions in stimulus processing during the FN, N2, and ERN (Aviyente et al., 2017; Cavanagh et al., 2009, 2014; Watts et al., 2018), where increases in ICPS are associated with increases in control functions.

Substantial empirical work has demonstrated a fundamental coupling between ACC and lateral PFC (lPFC; Duvernea and Koechlin, 2017; Miller and Cohen, 2001), in which the ACC regulates the engagement of the lateral PFC in cognitive control paradigms (Botvinick et al., 2001; Holroyd & Coles, 2002) and during reward/motivational incentives (Kouneiher, Charron, & Koechlin, 2009). Increased medial to lateral PFC theta phase synchrony has also been demonstrated using EEG phase synchrony measures, during trials which are conflicting, surprising, or represent a motivational incentive (Hanslmayr et al., 2007; Luft, 2014; Smith et al., 2015); during performance feedback tasks (Aviyente et al., 2017; Watts et al., 2018); and time estimation tasks (Van de Vijver et al., 2011).

### Summary and current aims

Substantial evidence now indicates that activity in the anterior cingulate cortex (ACC) is a consistent predictor of symptom severity and treatment outcomes and can be measured by medial frontal theta event related potentials (ERPs). However, ERPs have yet to be evaluated as predictors of treatment response. There is also a lack of research on the functional networks associated with ACC symptom change, despite implications for prefrontal engagement of cognitive control processes. Therefore, the present study aimed to examine these gaps in the literature by using task-based electroencephalography (EEG) and medial frontal theta activity (MFT) as predictors of response to intervention. The primary outcome of interest was change in anxiety sensitivity, which serves as a broad mediating factor relevant to many internalizing conditions as well as PTSD.

We assessed the amplitude and inter-channel phase synchrony of MFT (theta-FN, theta-N2) as predictors of intervention related change in anxiety sensitivity. We hypothesized that increased midfrontal theta-FN and theta-N2 amplitude is expected to predict greater treatment response. With respect to phase synchrony, the directionality of effects could not be predicted based on limited work in this area. We also evaluated whether the MFT components represent a shared or unique process in relation to treatment related response in anxiety sensitivity and we expected theta-FN and theta-N2 to have shared variance in predicting change.

## Methods

### Participants

The current analysis utilized unpublished data from 233 participants (129 females; M age= 35.03 years, SD= 15.91 years) from a study at Florida State University that assessed neurophysiological markers and psychological risk factors related to anxiety and depression. All participants were 18 years of age or older and were screened for neurological conditions, visual impairments, and/or traumatic brain injuries.

Eligibility for the study was based on an assessment of risk factors for anxiety and depression, as determined by elevated scores on the 18-item self-report Anxiety Sensitivity Index-3 (ASI-3; Taylor et al., 2007) or elevated scores on the 15-item self-report Interpersonal Needs Questionnaire (INQ-15; Hill & Petit, 2013; Van Orden, Cukrowicz, Witte, & Joiner, 2012). Individuals were excluded from participation if they met any of the following criteria: significant medical illness, current substance dependence, current or past psychotic spectrum disorder, uncontrollable bipolar disorder, or serious suicidal intent.

### Intervention procedures

Participants were divided into four groups: one control group, which received weekly suicide risk evaluations by phone, and three treatment groups, which received interventions for anxiety, depression or both. Each participants of the control group completed a baseline neurophysiological evaluation with their personally-assigned study coordinator and was subsequently contacted by them once per week for a suicide risk evaluation. The group that received the anxiety interventions (Anx-RR) received a combination of Cognitive Anxiety Sensitivity Treatment (CAST) and Cognitive Bias Modification-Interpretation (CBM-I) for AS. The group that received the depression interventions (Mood-RR) received a CBM-I for Mood intervention and psychoeducation. Finally, the combined group received all the interventions above.

#### Anxiety Intervention Condition (ANX-RR)/ CAST Group

CAST is a fully computerized, 45-minute intervention designed to model the techniques, both educational and behavioral, that are commonly used in anxiety treatments. It aims to teach participants that the physiological arousal associated with stress is non-threatening. First, participants were directed to complete a standardized assessment of their fear to different arousal sensations. Then, they completed exposure trials of engaging in an arousing sensation (i.e., hyperventilation) followed by rating the level of arousal they experienced (scale of 1 to 10, with 10 being the highest). They were told that they would repeat each exercise until their subjective rating of distress was rated as minimal (0-1). They were also instructed to complete one set of each of the exercises daily until none of the exercises generated any fear/distress.

During the CBM-I task, participants were presented with a word (e.g., “excited”) for 1 second followed by a sentence (e.g., “You notice your heart is beating faster”), and asked to determine if the pairing was related. Half the time, word-sentence combinations created a benign meaning (previous example), while the other half of trials created an anxious meaning (e.g., “stressful” followed by “Your mind is full of thoughts”). Correct answers determined the anxious combinations to be “unrelated” and the benign combinations to be “related”. Participants received feedback on their answers and incorrect answers triggered an additional horn blast (approximately 85 decibels). Participants completed 40 test trials with no reinforcement (feedback), followed by 80 training trials with feedback/reinforced. Participants then took a 5-minute break during which they completed a filler task (simple math problems), followed by another 80 training trials. At the end, they were given 40 test trials of novel words and sentences that they had not seen before.

#### Mood Intervention Condition (MOOD)/ CBM-I Group

The mood condition paralleled the anxiety condition in that it included the psychoeducational and CBM-I portions, however the techniques being modeled are those that treat mood disorders. Participants completed a computerized, 50-minute intervention designed to model the techniques, both educational and behavioral, that are commonly used in mood disorders treatment. In the mood intervention, participants were taught that negative beliefs about being isolated and burdensome are usually inaccurate.

#### Repeated Contact Control Condition (RCC)/ Control Group

Participants in the RCC condition were assigned a personal study coordinator with whom they met once per week for three weeks (corresponding to the treatment session intervals for those in the active treatment conditions) for a brief check-in where suicide risk was evaluated and preventative measures were taken if needed (e.g., safety plan, lethal means counseling, resource recommendations).

Response data were not collected in this dataset, so the effectiveness of each treatment component (Anx-RR v. Mood-RR v. psychoeducation) is unknown. However, the cumulative effectiveness of the intervention can be measured by examining change in symptom severity compared to the control group.

### Measures and tasks

This study uses EEG to evaluate MFT time frequency amplitude and inter-channel phase synchrony as predictors of treatment response. These measures are rarely studied, especially in relation to treatment response, making this approach uniquely valuable. Participants also completed the Beck Anxiety Inventory (BAI; Steer and Beck, 1997), as well as the Beck Depression Inventory-II (BDI-II; Beck, Steer, & Brown, 1996). Participants also completed a series of tasks designed to assess cognitive and affective processes (e.g., emotional picture viewing tasks, visual oddball task, gambling feedback, go/no-go, resting state). The current analyses focused on the gambling and go/no-go tasks since these are reliably used in generating the FN (gambling feedback) and N2 (go/no-go) ERPs.

The gambling task was a modified version used by Gehring and Willoughby (2002) in which the participant chose between two monetary options on each trial and then received feedback indicating whether the choice resulted in winning or losing money on that trial. After subjects made a choice by pressing the left or right button, the chosen box turned either red (loss) or green (win). This feedback stimulus was presented 1000 ms after the button press and appeared for 1000 ms, followed by a blank screen for 1500 ms preceding the onset of the next trial. Based on the design used by Gehring and Willoughby (2002), all four possible combinations of 5 and 25 (i.e., 5-5, 5-25, 25-5, and 25-25) were evenly crossed with the four possible win/loss outcomes (win-win, win-loss, loss-win, loss-loss), resulting in 16 trial types. Stimulus combinations and win/loss feedback on all trials were randomly determined, such that future trials were not predictable from outcomes associated with prior choices. Two sets of these 16 trial types, ordered randomly, were included in each block. Upon completion of a block, participants received feedback about their win/loss ratio within that block. Participants completed 7 blocks of 32 trials each.

For the Go/No-go task, participants were presented with two different white letters (e.g., S-F) displayed sequentially on a black background, and instructed to press the right or left button corresponding to the letter that appeared (go trials; e.g., S=left, F=right). However, when the stimulus repeated itself participants were asked to withhold their response (no-go trials; e.g., the third letter in the string S-F-F-S-F). No-go trials were pseudo-randomly interspersed throughout the task such that one, three, or five go trials always preceded a no-go trial. Seven blocks of twenty-four trials each were completed, with eighteen go trials (75%) and six no-go trials (25%) in each block. Stimulus duration was 296 ms, the response window was 1150 ms, and the inter-trial-interval was 900 ms. Participants also completed a practice version of the task consisting of 20 practice trials with different letters, but no electrophysiology data was recorded during the practice. Participants were provided with their overall accuracy after each block, and associations between the letter (e.g., S or F) and right/left button presses were counterbalanced across participants.

### Psychophysiological data acquisition & processing

All neurophysiological data was collected in a dimly lit sound attenuated room, where E-prime version 2.0 was used to present the computer tasks. Experimental stimuli was presented on a 21-inch Dell high definition CRT color monitor, centrally placed in front of participants at a viewing distance of 100 cm.

Neurophysiological data was recorded using a BrainVision 96-channel actiCap (sintered Ag-Ag/Cl; international 10-20 system) as well as a 24-bit battery-supplied active channel amplifier. Horizontal electrooculogram activity was recorded from electrodes placed on the outer canthus of both eyes, while vertical electrooculogram activity was recorded from electrodes placed above and below the left eye. Impedances were kept below 10 kΩ. EEG signals were vertex referenced during recording, and referenced to average mastoid signals offline (electrodes TP9 and TP10). Recordings were collected using a 500Hz sampling rate, analog 0.05 to 100Hz bandpass filter, and digitized at 1000 Hz using BrainVision PyCorder (Brain Vision LLC).

Epochs of three seconds were taken from 1000 ms pre to 2000 ms post stimulus with a 150 ms to 10 ms pre-stimulus baseline, and re-referenced to averaged mastoid sites. Data was corrected for ocular artifacts using an algorithm developed by Semlitsch and colleagues (1986) in the Neuroscan Edit 4.5 software (Neuroscan, Inc.), and downsampled to 128 Hz using the Matlab resample function (Mathworks, Inc.), which applies an anti-aliasing filter during resampling. Then, two methods of data cleaning were used. In the first method, trials were rejected if activity at frontal electrodes (F3 or F4) exceeded ±100 µV in either the pre-stimulus period of -1000 to -1 ms, or the post stimulus period of 1 to 2000ms. Within-trial individual electrodes were rejected if activity exceeded ±100 µV during the same pre- and post-stimulus time periods. This removed 14.9% of all trials from the gambling task, and 13.8% of all trials from the Go/No-go task. Additionally, visual analysis of the averaged waveforms indicated that 94 electrodes out of 25,850 (0.4%) were disconnected during recording in the gambling task, and 135 (0.5%) were disconnected during recording in the Go/No-go task. These electrodes were replaced with the mean of the three nearest neighbors. After preprocessing, the data were averaged according to each stimulus category for N2 (i.e., go, no-go) or feedback category for FN (i.e., gain, loss).

### Time-frequency amplitude

Time-frequency principal components analysis (TF) analysis is a technique that can be used to disentangle overlapping spatial and temporal frequency band effects in several common ERP signals, including the FN and N2 components (Bernat, Williams, & Gehring, 2005; Bernat, Malone, Patrick, & Iacono, 2007; Bernat, Nelson, & Baskin-Sommers, 2015; Harper, Malone, & Bernat, 2014). The method used to isolate theta-N2 from theta-FN was identical to that of Bernat et al. (2011) and Nelson et al. (2011). Specifically, the condition-averaged stimulus-locked N2 and feedback-locked FN ERP signals were filtered using a 2 Hz highpass and 7 Hz lowpass Butterworth filter (all filters implemented with the matlab butter and filtfilt functions, matlab version 7.4, Mathworks, Inc.) as apart of a theta-filtered TF-PCA. In each PC, activity was appropriately isolated to best represent theta-N2 and theta-FN. Next, the theta-filtered signals were transformed into time-frequency energy representations using the binomial reduced interference distribution (RID) variant of Cohen’s class of time-frequency transforms.

For each of these TF transforms, principal components analysis was applied to an area corresponding to the 0 to 1000 ms time range (post-feedback for theta-FN, post-stimulus for theta-N2, respectively) and 0 to 14 Hz frequency range; this yielded equivalent time windows for decomposition, but with filters having narrowed the frequency activity within the window to theta, as described above. Principal components analysis (PCA) was used to identify the primary activation component in each ERP signal (theta-FN and theta-N2), corresponding to the largest principal component emerging from the principal components analysis (i.e., the component accounting for the greatest proportion of shared covariance across all time-frequency points). Details for this application of PCA to time-frequency surfaces have been previously published (Bernat, et al., 2005). Briefly, this involves first vectorizing the time-frequency surfaces (e.g., concatenating each frequency row end to end) such that the columns of the data matrix index data from different time-frequency points while the rows index the condition averages (separated by subject and electrode). The PCA decomposition is then conducted using this matrix, utilizing the covariance approach and varimax rotation. The vectorized components are then reassembled into time-frequency surface matrices for interpretation.

Figure 1 displays the grand average TF-PCA decomposition for theta in the Gambling (left) and Go/No-go (right) tasks. For both the Gambling and Gonogo tasks, two component TF-PCA solutions were extracted. In both solutions, tThe variance accounted for by the first two principal components (PCs) far exceeded that accounted for by the next PC (e.g., PC3= 6.52% and 8.31%, and PC4= 5.79% and 5.68%, respectively), indicating that retention of two PCs was justifiable. For the Gambling task,both PCs accounted for a total of 43.59% of the variance; for Gonogo, 51.47%. N1/P2 activity was best characterized in TF-PC1, accounting for 30.85% of the variance, while FN activity was best characterized in TF-PC2,accounting for 12.74% of the variance. In Gonogo, N1/P2 activity was best characterized in TF-PC1, accounting for 34.65% of the variance. The N2 was best characterized in TF-PC2, and accounted for 16.81% of the variance. PCs during the FN window (Gambling-PC2) and N2 window (Go/No-go-PC2) served as the primary variables of interest. Electrode FCZ was most proximal topographically to the center of activation for theta-FN during gain and loss conditions as well as theta-N2 during go and no-go conditions. Therefore, the average of three electrodes in medial frontal (FCZ, FC1, FC2) areas will be included in analyses.

**Figure 1.**
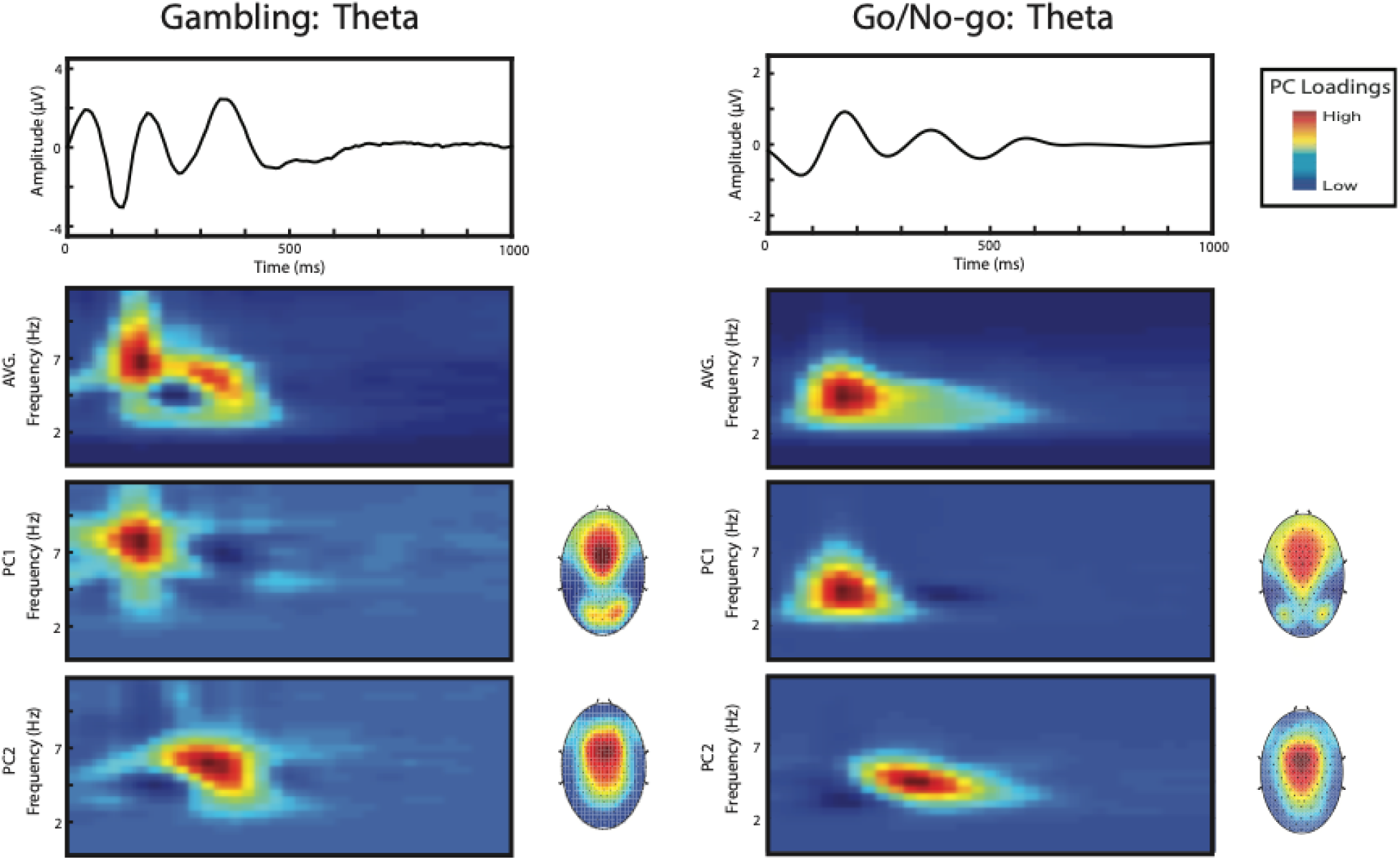
Grand average time-frequency (TF) decomposition of theta during the Gambling task (left) and the Go/No-go task (right) using principal components analysis (PCA) across all trial types (i.e., gain and loss trials in Gambling, & go and no-go trials in Go/No-go). *Waveform plots, top level*: Average time-domain ERPs across all trials, theta frequency-filtered. *Color surface plots, second level*: grand average time-frequency effects, with principal components depicted beneath. *Colored topographical head maps*: scalp topography distributions for the mean of the respective principal components. Gambling theta effects are best captured by two principle components, where PC1 represents theta-N1/P2, and PC2 represents theta-FN and is maximal at FCZ regions. Go/No-go theta effects are also indicated by two principal components, where PC1 represents N1/P2, while PC2 represents N2 and is maximal at FCZ regions.

Inter-channel phase synchrony (ICPS) indexes the degree of phase alignment between two electrode sites, and can be considered an index of functional connectivity between regions (Cavanagh et al., 2009; Cohen, 2011). Medial frontal theta components were assessed for phase synchrony with lateral prefrontal electrodes (Figure 2) over the FN and N2 windows. To avoid spurious phase synchrony between scalp electrodes due to volume conduction (Srinivasan et al., 2007), all EEG epochs were first transformed using current source density (CSD), which minimizes volume conduction by source localizing activity toward the cortical surface (Tenke and Kayser, 2012). Subsequently, phase-synchrony was computed as a phase locking value (PLV; Aviyente et al., 2011; Lachaux et al., 1999), used to quantify the synchrony between medial frontal and bilateral frontal theta EEG signals, separately for the FN and N2 windows. The mean of each bilateral electrode pair was used (i.e., mean of FCZ – F3 and FCZ – F4 PLVs). The PLV values were used to predict magnitude of treatment response at mid-treatment, 1-week post treatment, and 6 months post treatment.

**Figure 2.**
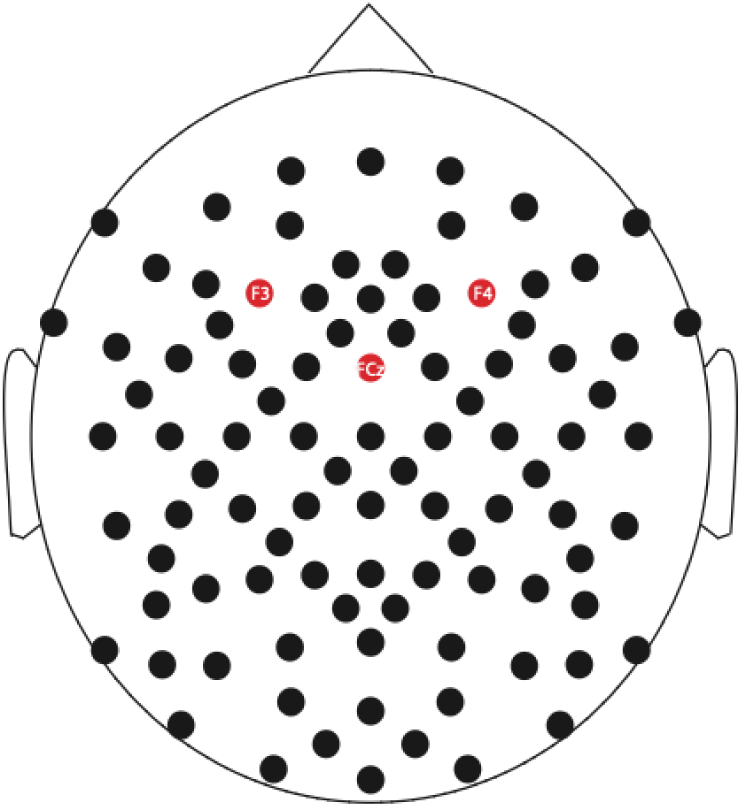
Topographical representation of electrodes in the 96-channel EEG cap. Red indicates medial frontal (FCZ) and lateral frontal (F3 & F4) electrodes included in ICPS analyses for theta-FN and theta-N2.

### Data analytic methods

Primary symptom change analyses were conducted in two steps. Step 1 involved reducing the number of bivariate comparisons and to represent an ‘omnibus’ model where all variables of interest were considered simultaneously. Linear Mixed Models were run separately for each neurophysiological predictor (theta-FN gain, theta-FN loss, theta-N2 go, theta-N2 no-go) for both amplitude and inter-channel phase synchrony, resulting in 8 models. In all models, separate terms were included for each of the treatment groups and dummy coded relative to the control. For simplicity, groups were labeled as Group1 (combined treatment), Group2 (Anx-RR), and Group3 (Mood-RR), with the intercept as the control group. Terms of interest within each model include the three Group x MFT interactions (e.g., Group1 x Theta-FN gain amplitude; Group2 x Theta-FN gain amplitude; Group3 x Theta-FN gain amplitude), as well as the three Group x MFT x Time interactions. Within each model, the two-way interactions test whether the MFT variable differentially predicts treatment response based on treatment group. The three-way interactions with time tested whether this prediction effect differs as a function of time. An unstructured covariance matrix with restricted maximum likelihood estimation was used. Due to the number of models being tested, Bonferroni p-value correction was applied such that significance in the models is based on p<.0125 (.05/4 for amplitude and .05/4 for ICPS).

Step 2 of the analyses involved specific effects testing with the purpose of assessing the specificity, sensitivity, and incremental validity of any significant effects identified in step 1. To do this, significant interactions from the linear mixed models were further assessed with nonparametric spearman correlations. Specificity is indicated here by effects that are present for one group and not the other/s. Sensitivity is demonstrated by reliability (i.e., consistency across time) and accuracy (i.e., percent of individuals classified as responders versus non-responders). To test for incremental validity, partial spearman correlation coefficients are reported after controlling for baseline symptom severity, and other self-report measures (including anxiety and depression) that may be quicker and more cost-effective predictors. Age and gender were also assessed in relation to treatment response. Multiple comparison correction was applied based on the number of correlations performed.

Finally, theta-FN and theta-N2 were assessed for unique versus shared effects in relation to treatment outcome by entering both into a regression model predicting treatment response.

For demographic purposes and to further illustrate treatment effectiveness, subjects will be characterized as achieving (1) a full response (≥50% improvement in ASI scores from baseline), (2) a partial response (<50% but ≥25% improvement), or (3) no response (<25% improvement), based on thresholds used in other treatment response studies (Nierenberg & DeCecco, 2001). Chi-square tests and Fisher’s Exact Tests will be used to compare response rates between treatment and control groups, and between ‘responders’ (≥50% improvement) versus ‘non-responders’ (<50% improvement).

## Results

### Behavioral results

Reaction times and accuracies were computed for the Go and No-go trial types. Correct responses to Go stimuli had a mean reaction time (RT) of 586.57 ms (SD= 138.93), and incorrect responses to No-go stimuli (false alarms) had a mean RT of 1969.08 ms (SD= 239.27). Mean accuracy rates for Go trials was 94.94% (SD= 9.10), while the mean accuracy for No-go trials was 83.47% (SD= 14.58). The four treatment groups did not differ on mean RT to Go (p=.802) or No-go (p=.672). There were also no significant differences between groups on accuracy for Go trials (p=.355) or No-go trials (p=.708), indicating the four treatment groups did not differ in their behavioral performance. Reaction time and accuracy was not applicable in the gambling feedback task.

### Demographics & treatment efficacy

Demographic characteristics for all four groups of subjects as well as their baseline severity scores (i.e., ASI) and response rates are shown in Table 1. The four groups do not significantly differ on age (p=.449), gender (p=.536), race (p=.292), or psychopathology (p=.356), indicating these factors were reasonably balanced across groups.

**Table 1.** Demographics and outcome indices for individuals in the treatment and control groups.

|  | **Combined**<br>**(N=60)** | **Anx-RR**<br>**(N=60)** | **Mood-RR**<br>**(N=57)** | **Control**<br>**(N=56)** |
| --- | --- | --- | --- | --- |
| **Age (mean years, SD)** | 35.2(15.04) | 34.2(16.76) | 34.1(15.58) | 36.7(16.49) |
| **Gender (N female)** | 29(48%) | 37(62%) | 32(56%) | 31(55%) |
| **Race (N)** |  |  |  |  |
| white | 37(61%) | 38(63%) | 35(61%) | 31(55%) |
| black | 14(23%) | 12(20%) | 13(23%) | 20(35%) |
| other | 9(15%) | 10(17%) | 9(16%) | 5(9%) |
| **Psychopathology (N)** |  |  |  |  |
| Anxiety disorders | 20(33.3%) | 29(48.3%) | 19(33.3%) | 24(42.8%) |
| Depressive disorders | 13(21.7%) | 12(20%) | 16(28.1%) | 9(16.1%) |
| Bipolar disorders | 1(1.7%) | 2(3.3%) | 1(1.7%) | 2(3.6%) |
| Trauma disorders | 10(16.7%) | 9(15%) | 13(22.8%) | 6(10.7%) |
| Obsessive compulsive disorders | 2(3.3%) | 2(3.3%) | 1(1.7%) | 4(7.1%) |
| Substance use disorders | 4(6.7%) | 0(0%) | 1(1.7%) | 1(1.8%) |
| Other (Axis II, somatic) | 2(3.3%) | 1(1.6%) | 2(3.5%) | 3(5.4%) |
| No diagnosis | 8(13.3%) | 5(8.3%) | 4(7%) | 7(12.5%) |
| **Baseline severity**<br>**(mean ASI, SD)** | 32.7 (16.8) | 32.3 (17.4) | 28.3 (16.0) | 32.5 (17.7) |
| **Post-Treatment response rates** |  |  |  |  |
| Responder ($\geq 50\%$ reduction) | 38/55 (69.1%) | 32/59 (54.2%) | 16/54 (29.6%) | 18/54 (33.3%) |
| Partial Response (25-49% reduction) | 10/55 (18.2%) | 17/59 (28.8%) | 17/54 (31.5%) | 12/54 (22.2%) |
| No Response ($< 25\%$ reduction) | 7/55 (12.7%) | 10/59 (16.9%) | 21/54 (38.9%) | 24/54 (44.4%) |

Importantly, however, the treatment and control groups do show significant differences in response rates (Table 1, χ2 =29.13, df=6, p<.001). As shown in Figure 3, by one week post-treatment the combined treatment group had the highest percent improvement (M=63.77, SE=3.97), which was significantly greater than the Anx-RR group (M=45.43, SE=5.78; t=2.58, p=.011), the Mood-RR group (M=27.23, SE=6.46; t=4.82, p<.001), and the control group (M=15.50, SE=9.74; t=4.59, p<.001). The Anx-RR group had the second highest percent improvement, which was significantly greater than the Mood-RR group (t=2.1, p=.038) and the control group (t=2.7, p=.008). The Mood-RR and control groups did not differ in treatment response magnitudes (t=1.0, p=.318).

**Figure 3.**
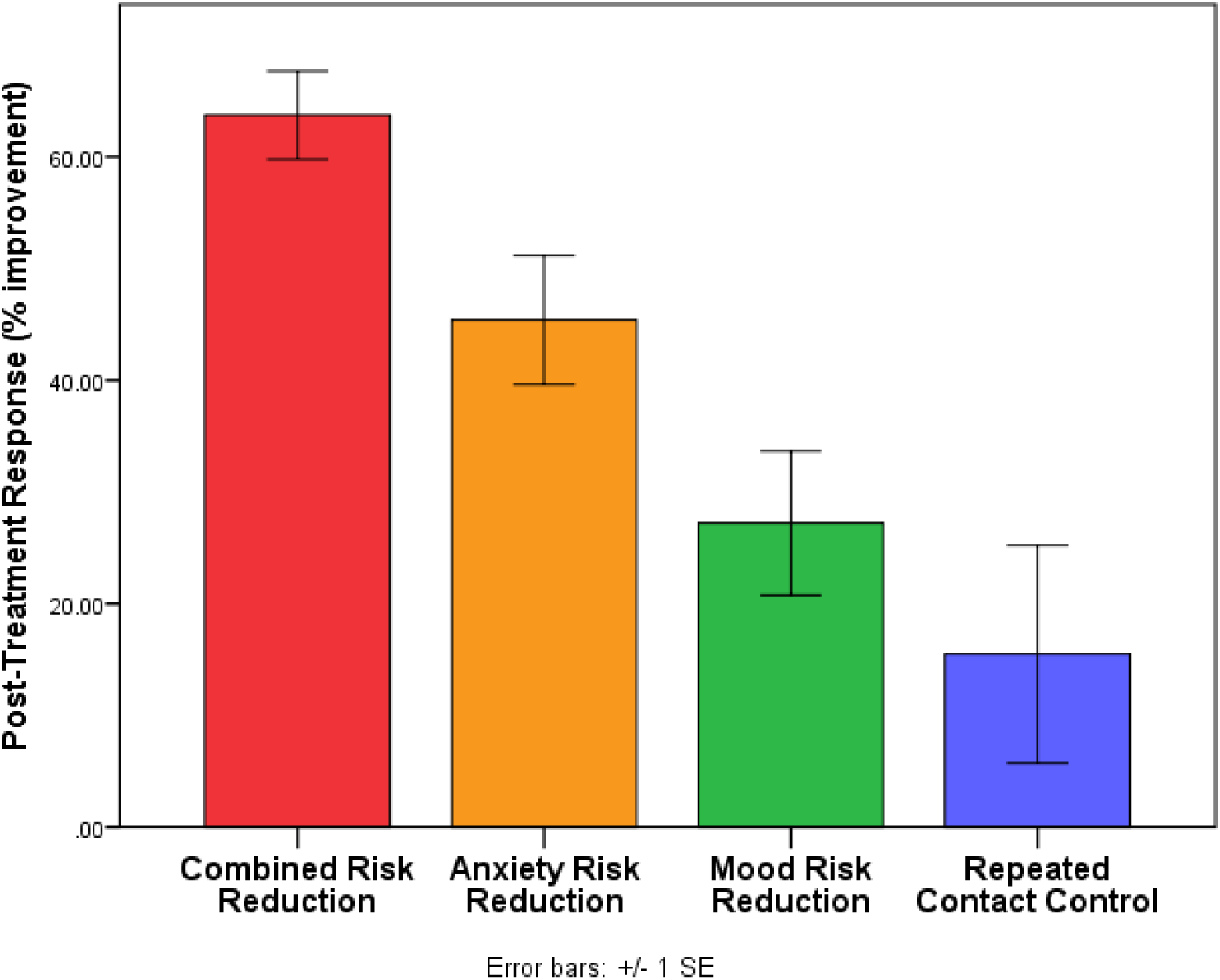
Post-treatment (1 week post) response rates according to intervention group. Individuals in the Combined-RR group had significantly greater improvement in anxiety sensitivity symptoms compared to all other groups. The Mood-RR/CBM-I group did not significantly differ from the control group.

The combined treatment group also had a significantly greater proportion of individuals achieving full response (>50% improvement) compared to the control group (χ2 =13.85, p<.001). Specifically, 38 of 55 individuals (69.1%) in the treatment group achieved full response (Table 1), compared to 18 of 54 (33.3%) in the control group. Additionally, only 7 individuals in the combined treatment group (12.7%) experienced no response, while almost half of the control group (24/54; 44.4%) failed to show a response. This difference was also statistically significant (χ2 =13.34, p<.001), indicating the combined treatment was the most effective method of reducing anxiety sensitivity, compared to a repeated contact control condition.

In addition to 1 week post-treatment, treatment effects are also apparent at the mid-treatment time point as well as 6 months post-treatment (Figure 4). As shown in Figure 4, the combined treatment group had significantly lower anxiety sensitivity scores at mid-treatment and 1-week post treatment compared to the control group (p’s<.001). This difference in symptom severity continued at trend level by 6 months post-treatment. Importantly, the groups did not differ in baseline severity

**Figure 4.**
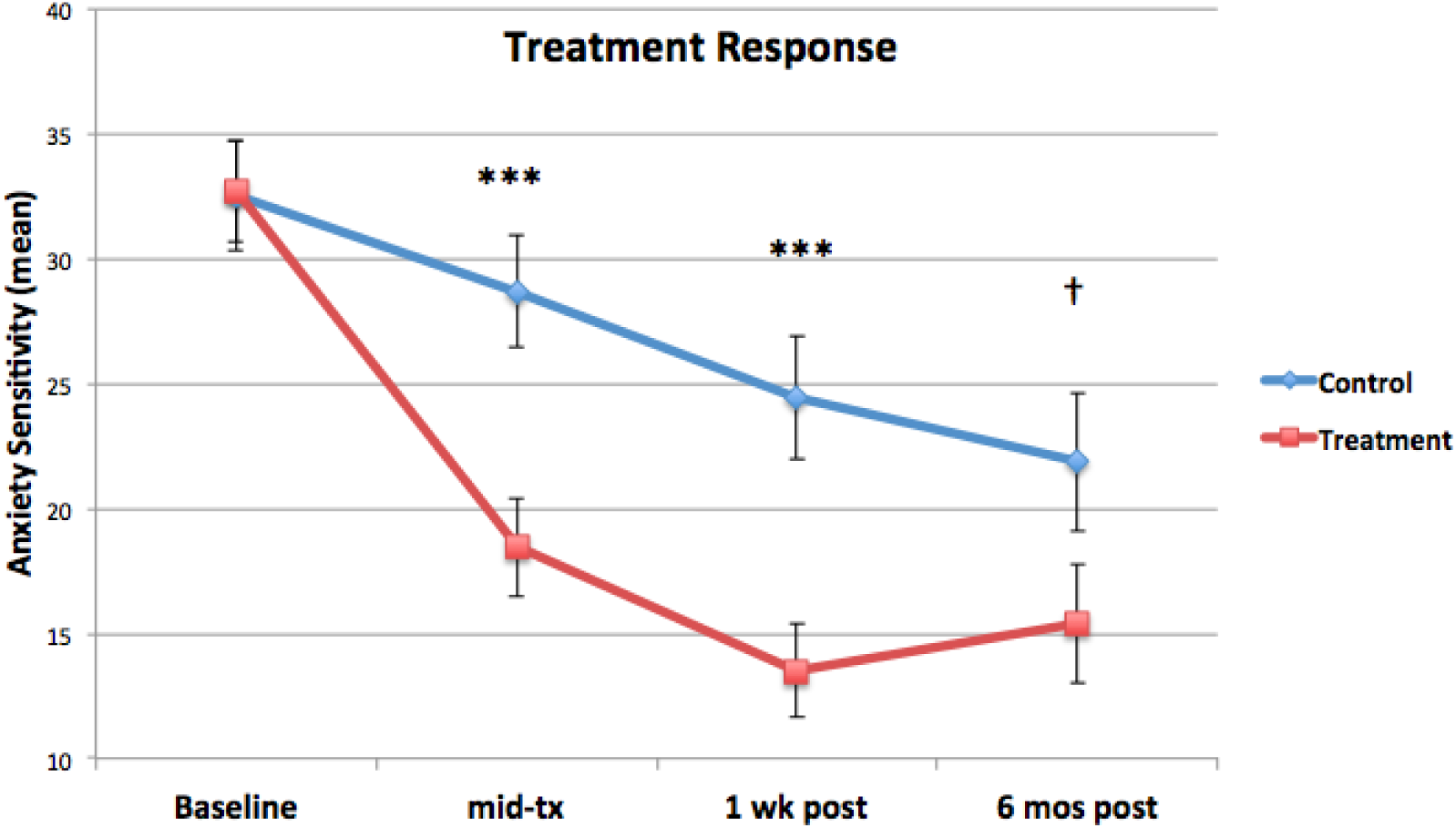
Change in symptom severity over time between the control (blue) and Combined treatment (red) groups. Symptom severity was defined by mean scores on a continuous measure of anxiety sensitivity (AS). Improvement in symptoms is indicated here by a reduction in mean AS scores. Statistical comparisons indicate significant differences in symptom reduction between the control and treatment groups at mid-treatment and 1 week post-treatment, with a trend level difference at 6 months post-treatment. The groups did not differ at baseline. Note: ***p<.001, † p<.10

### Neurophysiological data

A one-sample t-test indicated the loss – gain difference for FN to be significantly less than zero (M= -1.72, SD= 2.36 μV; t= -11.27, p<.0001). Additionally, the loss-gain FN difference in theta was significantly greater than zero (M= .10, SD= .157; t= 10.02, p<.0001). Similarly for the N2, no-go – go differences in theta were significantly greater than zero (M= .05, SD=.08; t=9.45, p<.0001).

### Predictors of treatment response: Step 1, Linear Mixed Models

#### Model 1: Theta-FN Gain

Results from the first linear mixed model show a non-significant Group1 x Theta-FN Gain interaction on anxiety sensitivity severity (F=.378, df=283.41, p=.539), indicating that Theta-FN amplitude to gains did not differentiate the combined treatment and control groups on treatment response. Within the same model, there was a non-significant Group2 x Theta-FN Gain interaction (F=.852, df=282.12, p=.357), as well as a non-significant Group3 x Theta-FN Gain interaction (F=.196, df=284.15, p=.659). These results show that Theta-FN Gain amplitude did not predict treatment response in the combined, Mood-RR, or Anx-RR treatment groups relative to the control group.

These relationships were also tested in a three-way interaction with time in the model. All three tests were non-significant, including Group1 x Theta-FN Gain x Time (F=.093, df=418.03, p=.760), Group2 x Theta-FN Gain x Time (F=.671, df=411.37, p=.413), as well as Group3 x Theta-FN Gain x Time (F=1.72, df=422.48, p=.190).

#### Model 2: Theta-FN Loss

Similarly for Theta-FN Loss amplitude, there was a non-significant Group1 x Theta-FN Loss interaction on anxiety sensitivity severity (F=2.32, df=284.39, p=.128), as well as non-significant interactions for Group2 x Theta-FN Loss (F=.032, df=282.35, p=.858), and Group3 x Theta-FN Loss (F=.015, df=284.45, p=.901). These results indicate that Theta-FN Loss amplitude does not significantly predict treatment response to the combined, Anx-RR, or Mood-RR groups, relative to control.

These effects were also not modulated by time as shown by non-significant three-way interactions for Group1 x Theta-FN Loss x Time (F=.403, df=421.59, p=.526), as well as Group2 x Theta-FN Loss x Time (F=.011, df=410.56, p=.916), and Group3 x Theta-FN Loss x Time (F=2.51, df=421.90, p=.114).

#### Model 3: Theta-N2 Go

Results indicate non-significant two-way interactions for Group1 x Theta-N2 Go (F=1.48, df=270.74, p=.224), as well as Group2 x Theta-N2 Go (F=.114, df=270.17, p=.736), and Group3 x Theta-N2 Go (F=.593, df=270.23, p=.442). Similar to the above models, these results indicate that Theta-N2 Go amplitude does not significantly predict treatment response to the combined, Anx-RR, or Mood-RR groups relative to control.

The three-way interactions with time were also non-significant for each of these terms, including Group1 x Theta-N2 Go x Time (F=.917, df=405.16, p=.339), Group2 x Theta-N2 Go x Time (F=.204, df=402.49, p=.652), and Group3 x Theta-N2 Go x Time (F=.080, df=402.88, p=.777).

#### Model 4: Theta-N2 No-go

Similarly for Theta-N2 No-go amplitude, results show a non-significant two-way interaction for Group1 x Theta-N2 No-go (F=.278, df=270.15, p=.598), as well as Group2 x Theta-N2 No-go (F=.133, df=269.89, p=.715), and Group3 x Theta-N2 No-go (F=.533, df=269.63, p=.466). These results show that Theta-N2 No-go amplitude does not significantly predict treatment response in the combined, Anx-RR, or Mood-RR groups relative to control.

These effects were also not modulated by time, as shown by non-significant three-way interactions for Group1 x Theta-N2 No-go x Time (F=.003, df=405.34, p=.955), as well as Group2 x Theta-N2 No-go x Time (F=.487, df=403.81, p=.486), and Group3 x Theta-N2 No-go x Time (F=.848, df=402.72, p=.358).

### Inter-channel Phase Synchrony (ICPS)

#### Model 1: ICPS Theta-FN Gain

Results from the first model indicate a significant two-way interaction for Group1 x ICPS Theta-FN Gain (F=6.98, df=285.99, p=.009), indicating that Theta-FN phase synchrony significantly predicts treatment response to the combined group relative to the control. The three-way interaction for Group1 x ICPS Theta-FN Gain x Time was non-significant (F=2.07, df=425.92, p=.150), indicating that the above treatment response effect does not differ as a function of time.

The other treatment groups showed non-significant two-way interactions for Group2 x ICPS Theta-FN Gain (F=1.56, df=286.33, p=.213), as well as Group3 x ICPS Theta-FN Gain (F=2.71, df=290.29, p=.101), indicating that ICPS Theta-FN Gain does not predict treatment response to the Anx-RR or Mood-RR treatment groups relative to the control group. Finally, there were no significant three-way interactions for Group2 x ICPS Theta-FN Gain x Time (F=.006, df=417.34, p=.941) or Group3 x ICPS Theta-FN Gain x Time (F=2.09, df=466.19, p=.148).

#### Model 2: ICPS Theta-FN Loss

Similarly for ICPS Theta-FN Loss, there was a significant two-way interaction for Group1 x ICPS Theta-FN Loss on treatment response (F=10.82, df=286.39, p=.001), indicating that Theta-FN Loss phase synchrony predicts treatment response to the combined group relative to the control group. This effect was not modulated by time as shown by a non-significant three-way interaction between Group1 x ICPS Theta-FN Loss x Time (F=.165, df=413.88, p=.685).

ICPS Theta-FN Loss did not predict treatment response to the Anx-RR or Mood-RR groups relative to control, as shown by a non-significant Group2 x ICPS Theta-FN Loss (F=1.89, df=286.26, p=.170) and Group3 x ICPS Theta-FN Loss (F=3.29, df=291.39, p=.071) interactions. Three-way interactions for these terms were also non-significant (Group2 x ICPS Theta-FN Loss x Time: F=.916, df=413.13, p=.339; Group3 x ICPS Theta-FN Loss x Time: F=1.637, df=453.17, p=.201).

#### Model 3: ICPS Theta-N2 Go

Results show a significant Group1 x ICPS Theta-N2 Go interaction (F=12.37, df=279.47, p=.001), indicating that Theta-N2 Go phase synchrony significantly predicts treatment response to the combined group relative to the control. This effect was not modulated by time, as shown by a non-significant three-way interaction (Group1 x ICPS Theta-N2 Go x Time: F=.422, df=411.83, p=.516).

Similar to the previous models, ICPS Theta-N2 Go did not predict treatment response to the Anx-RR or Mood-RR groups relative to the control (Group2 x ICPS Theta-N2 Go: F=.319, df=278.93, p=.573; Group3 x ICPS Theta-N2 Go: F=.110, df=283.62, p=.741). The three-way interaction for these terms was also not significant (Group2 x ICPS Theta-N2 Go x Time: F=.036, df=409.91, p=.851; Group3 x ICPS Theta-N2 Go x Time: F=2.80, df=432.07, p=.095).

#### Model 4: ICPS Theta-N2 No-go

Unlike the previous models, ICPS Theta-N2 No-go did not predict treatment response to the combined group (Group1 x ICPS Theta-N2 No-go: F=2.05, df=275.55, p=.153). It was also non-significant in predicting treatment response for the Anx-RR and Mood-RR groups relative to the control (Group2 x ICPS Theta-N2 No-go: F=.001, df=275.22, p=.971; Group3 x ICPS Theta-N2 No-go: F=.378, df=278.81, p=.539). Finally, similar to the above models, the three-way interactions for these terms did not reach threshold for significance (Group1 x ICPS Theta-N2 No-go x Time: F=.041, df=408.16, p=.840; Group2 x ICPS Theta-N2 No-go x Time: F=.868, df=407.15, p=.352; Group3 x ICPS Theta-N2 No-go x Time: F=4.69, df=425.67, p=.031).

### Predictors of treatment response: Step 2, Correlations

Spearman correlation coefficients were computed for the three ICPS variables (Theta-FN gain, Theta-FN loss, Theta-N2 Go) and treatment groups (Combined treatment v. control group) that demonstrated a significant relationship to predicting treatment outcomes. Based on the number of tests, multiple comparison corrected was applied such that significance is based on p<.0055 (.05/9). As shown in Table 2, lower pre-treatment ICPS Theta-FN to gains is significantly correlated with better treatment outcomes. This effect was significant at the mid-treatment assessment (p=.001), as well as 1 week post-treatment (p=.003), and in a similar direction, though not significant, at 6 months post-treatment (p=.120). A similar finding occurred for ICPS Theta-N2 to Go stimuli, such that lower pre-treatment phase synchrony is correlated with better treatment response at mid-treatment (p=.001), and 1 week post-treatment (p=.004), and is in a similar direction, though not significant, 6 months after treatment (p=.235). Scatterplots for these findings are depicted in Figure 5 with the corresponding spearman rho effects. Finally, although lower ICPS Theta-FN to loss was also associated with better treatment outcomes, the effect was only trend level at mid-treatment (p=.051), and did not reach significance 1 week post-treatment (p=.134), or 6 months post-treatment (p=.265).

**Figure 5.**
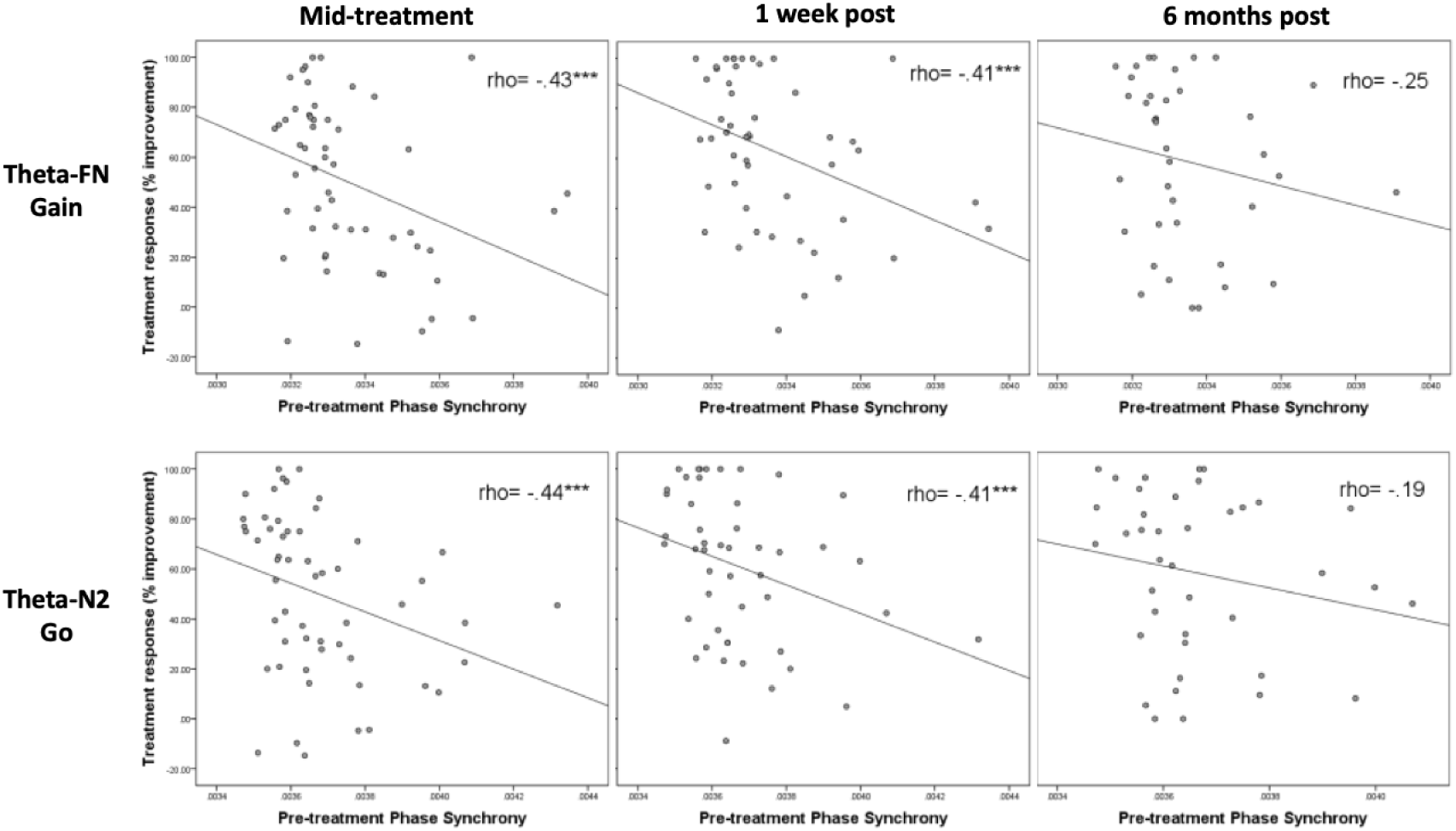
Scatterplots depicting relationships between pre-treatment phase synchrony for Theta-FN Gain and Theta-N2 Go with treatment response in the combined group at mid-treatment, 1 week post, and 6 months post-treatment

**Table 2.** Spearman correlations for pre-treatment ICPS with treatment response in the combined treatment group (top) and control (bottom).

|  |  | Treatment response: Combined group |  |  |
| --- | --- | --- | --- | --- |
|  |  | Mid-tx<br>(N=55) | 1 week post<br>(N=49) | 6 mos post<br>(N=39) |
| ICPS Theta-FN | Gain | -.43*** | -.41*** | -.26 |
|  | Loss | -.27<sup>†</sup> | -.22 | -.18 |
| ICPS Theta-N2 | Go | -.44*** | -.41*** | -.19 |
|  |  | Treatment response: Control group |  |  |
|  |  | Mid-tx<br>(N=52) | 1 week post<br>(N=47) | 6 mos post<br>(N=34) |
| ICPS Theta-FN | Gain | .22 | .16 | -.01 |
|  | Loss | .25<sup>†</sup> | .11 | .07 |
| ICPS Theta-N2 | Go | .01 | .03 | .03 |

Note: \*\*\*p<.005, <sup>†</sup>p<.1

### Specificity of treatment response

In contrast to the significant prediction effects for the combined treatment group, ICPS did not predict treatment outcomes to the control group (Table 2), thus demonstrating specificity of the treatment response effect. With the exception of ICPS Theta-FN to loss at trend level (p=.075), all other relationships did not approach significance in the control group (p>.10).

### Incremental validity: Examining potential covariates

Baseline symptom severity (i.e., mean ASI) as well as baseline depression and anxiety self-report measures were assessed as potential covariates. Baseline ASI significantly predicted treatment response at mid-treatment (p=.010), 1 week post-treatment (p=.021), as well as 6 months post-treatment (p=.010; Table 3). Baseline depression severity (BDI2) also demonstrated a significant relationship with treatment response at mid-treatment (p=.001), and 1 week post-treatment (p=.041), but was not significant at 6 months post treatment (p=.281; Table 3). Finally, baseline general worry (PSWQ) significantly predicted treatment response at all three assessment points (mid-treatment: p=.004, 1 week post: p=.002, 6 months post: p=.019; Table 3). As such, these variables were included as covariates in the analyses.

Age was not associated with treatment response at any of the assessment time points (mid-treatment: p=.791, 1 week post: p=.604, 6 months post: p=.835; Table 3). Gender was also not a significant predictor of treatment response at any of the assessments (mid-treatment: p=.733, 1 week post: p=.105, 6 months post: p=.908; Table 3)

### Incremental validity: Partial correlations

Partial spearman correlations were computed after adding baseline ASI severity, depression, and general worry as covariates. As shown in Table 4, results indicate ICPS Theta-FN to gains remains significant as a predictor of treatment response at mid-treatment (p=.001), 1 week post-treatment (p=.002), as well as 6 months after treatment has completed (p=.038). ICPS Theta-N2 to go’s also remains significant as a predictor of mid-treatment response (p=.030), but becomes non-significant at the 1 week post-treatment (p=.247) and 6 months post-treatment assessments (p=.801). Finally, ICPS Theta-FN to losses remains non-significant at all three assessment time points (mid-treatment: p=.158, 1 week post: p=.375, 6 months post: p=.708).

**Table 4.** Partial spearman correlations after including baseline severity, depression, and general worry as covariates.

|  |  | Treatment Response (% improvement) |  |  |
| --- | --- | --- | --- | --- |
|  |  | mid-tx<br>(N=55) | 1 week post<br>(N=49) | 6 mos post<br>(N=39) |
| ICPS<br>Theta-FN | Gain | **-.45\*\*** | **-.44\*\*** | **-.35\*** |
|  | Loss | -.20 | -.13 | -.07 |
| ICPS<br>Theta-N2 | Go | **-.30\*** | -.18 | -.04 |

Note: \*\* $p<.01$ , \* $p<.05$

### Sensitivity of treatment response: Accuracy

Given the findings above that low pre-treatment theta phase synchrony during gain and go trials predicts better treatment outcome, post-hoc sensitivity estimates were calculated. Using the median split, subjects were classified as ‘high’ or ‘low’ on ICPS Theta-FN Gain and ICPS Theta-N2 Go. Treatment response was also dichotomized into ‘Responder’ (>=50% improvement) and ‘Non-responder’ (<50% improvement) at 1 week post-treatment. As shown in Figure 6, 85% of individuals with low pre-treatment Theta-FN Gain phase synchrony were classified as responders, compared to 48% of individuals with high pre-treatment phase synchrony, a difference that was statistically significant (p=.013, Fisher’s Exact Test). A similar effect was found for pre-treatment Theta-N2 Go phase synchrony, where 85% of individuals with low phase synchrony were classified as responders, compared to 50% of individuals with high phase synchrony (p=.016, Fisher’s Exact Test)

**Figure 6.**
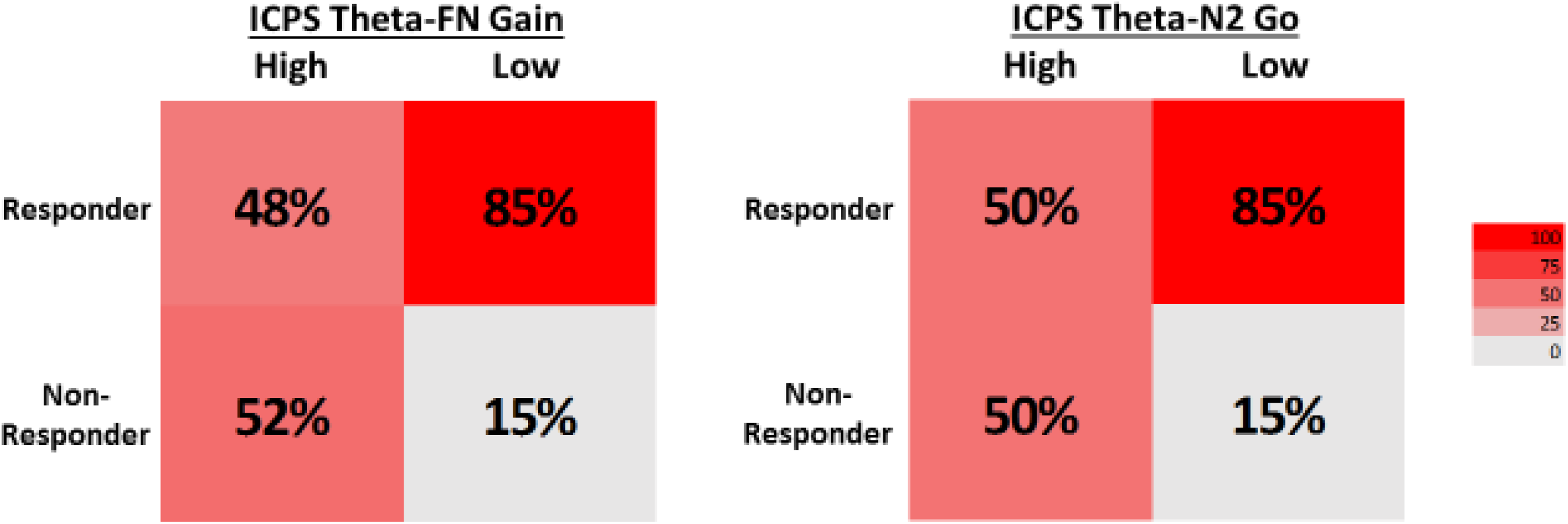
Percentage of individuals who are classified as a treatment responder versus non-responder at 1 week post-treatment, based on their high and low pre-treatment theta phase synchrony during gains (left) and go’s (right). Low theta phase synchrony at baseline differentiates responders from non-responders.

### Shared versus unique MFT effects

Given that ICPS Theta-FN Gain and ICPS Theta-N2 Go were both significant predictors of anxiety sensitivity treatment response, regressions were conducted to assess the unique versus shared contributions of each measure at all three assessment time points. As shown in Table 5, when both measures are entered into a multiple regression model, they each become non-significant in predicting treatment response, suggesting they have shared variance in relation to treatment outcomes. This shared variance effect was demonstrated at mid-treatment (Theta-FN Gain: p=.401, Theta-N2 Go: p=.317), as well as 1 week post-treatment (Theta-FN Gain: p=.530, Theta-N2 Go: p=.206), and 6 months post-treatment (Theta-FN Gain: p=.839, Theta-N2 Go: p=.292)

**Table 5.** Multiple regressions with ICPS predictors of treatment response.

|  | *B* | *t* | *p* | partial *r* | *R*<sup>2</sup> |
| --- | --- | --- | --- | --- | --- |
| 1. Mid-treatment response |  |  |  |  | .14 |
| ICPS theta-FN Gain | -.18 | -0.8 | .401 | -.12 |  |
| ICPS theta-N2 Go | -.22 | -1.0 | .317 | -.15 |  |
| 2. 1 week post-treatment |  |  |  |  | .16 |
| ICPS theta-FN Gain | -.14 | -0.6 | .530 | -.10 |  |
| ICPS theta-N2 Go | -.28 | -1.3 | .206 | -.19 |  |
| 3. 6 months post-treatment |  |  |  |  | .07 |
| ICPS theta-FN Gain | -.04 | -0.2 | .839 | -.04 |  |
| ICPS theta-N2 Go | -.23 | -1.1 | .292 | -.18 |  |

## Discussion

Previous research has demonstrated that the anterior cingulate cortex is a consistent and robust predictor of treatment response. While this effect has been shown across multiple interventions, clinical populations, and imaging modalities, it has not been assessed using task-based EEG paradigms (Banica et al., 2020; Cavanagh & Shackman, 2015; Ellis, Watts, Schmidt, & Bernat, 2018; Feurer et al. 2022; Hajcak 2008; Jaworska et al. 2023; Keedwell et al., 2010; McGrath et al., 2014; Meyer et al. 2015; Moser et al. 2013; Mueller et al., 2015; Pizzagalli et al. 2018; Saxena et al., 2003; Siegle et al., 2006, 2012; Spronk et al. 2011; Osinsky et al., 2017). EEG shows promise in clinical settings due to its greater temporal resolution and being a more portable and cost effective method of measuring brain activity compared to other imaging modalities. Despite these advantages and the ability to detect anterior cingulate activity through medial frontal theta ERPs (Banica et al., 2020; Cavanagh et al., 2012; Gehring & Willoughby, 2002; Hauser et al., 2014), these components have not been evaluated as predictors of treatment response. Further, there is a lack of research on the functional relationship between medial and lateral prefrontal regions, despite claims that these functional networks are implicated in treatment response. The current study therefore represents an initial study to assess task-based medial frontal theta amplitude and medial to lateral prefrontal theta phase synchrony as predictors of treatment response.

### Inter-channel Phase Synchrony (ICPS)

Phase synchrony results demonstrated significant prediction effects. ICPS Theta-FN Gain and ICPS Theta-N2 Go both significantly predicted treatment response, such that lower baseline medial to lateral prefrontal phase synchrony was associated with greater symptom improvement. This effect was specific to the gain and go conditions, as well as to the combined treatment group, relative to the CAST, CBM-I, and control treatment groups. As predictors, both ICPS variables demonstrated specificity, sensitivity, and incremental validity.

Regarding specificity (i.e., prediction to one group versus the other/s), symptom change was demonstrated for the combined treatment group only. The absence of MFT phase synchrony prediction response in the control group indicates the prediction effect is specific to the intervention and not symptom alleviation over time. Although the effect was in the same direction, MFT phase synchrony also did not predict in the CAST or CBM-I only treatment groups. This could be due to the combined treatment being the most effective and offering a longer intervention time since it combined both anxiety and depression components - relative to undergoing each component separately or not at all.

In addition to specificity, MFT phase synchrony also demonstrated *sensitivity* in the form of reliability/consistency and accuracy. Consistency of treatment response was demonstrated by a non-significant interaction with time in the linear mixed models - which indicates that the predictor maintained its predictive power over time - and by significant or trend level correlations at all assessments. Accuracy was demonstrated at 85% for low MFT phase synchrony when differentiating responders versus non-responders at the 1-week post treatment assessment - similar to other prediction estimates in the literature (85%; Furey et al., 2012). This effect maintained significance even after controlling for baseline severity and other self-report measures of depression and anxiety, demonstrating incremental validity. These are important indicators since consistency implies reliability and clinical utility as a marker of sustained symptom improvement, while accuracy indicates validity and MFT phase synchrony as a viable predictor of anxiety treatment outcomes. These highlight phase synchrony’s unique approach over self-report measures.

### Interpretation: Direction and condition-specific effects

Applying results from previous literature to the current findings, the low phase synchrony would indicate less engagement of cognitive control resources. Notably, heightened trait anxiety has been consistently linked to lower phase synchrony following errors in individual difference research (Barker et al., 2018; Comte et al., 2015; Moran et al., 2015) as well as the Klumpp and colleagues (2016) treatment response study. The authors examined predictors of CBT response in individuals with social anxiety disorder and, similar to the current study, found low functional connectivity predicted better outcomes. The authors concluded that low functional connectivity between medial and lateral PFC regions reflected poor regulatory ability, and suggested that these individuals may benefit the most from a treatment targeting the engagement of cognitive control processes. The authors claim that individuals with low prefrontal functional connectivity benefited more from the intervention because the cognitive behavioral treatment targeted the engagement of executive control processes. Specifically, it has been suggested that CBT draws upon executive functioning skills in a top-down manner (Bao et al., 2022; Mohlman & Gorman, 2005). Therefore, patients who have a deficit in this area may benefit more from the type of intervention that targets these cognitive processes. Although the current study does not employ therapist-delivered CBT, it utilizes cognitive behavioral techniques delivered online, therefore, it is possible that individuals with low phase synchrony between medial and lateral PFC are benefiting more from this particular treatment because components of the intervention engage cognitive control and regulatory processes in an effort to reduce anxiety.

Secondly, one additional area that requires further interpretation is why the ICPS prediction effects were strongest for the gain and go conditions, as opposed to the loss and no-go conditions. Evidence from basic science, individual difference research, and treatment outcome work points to a plausible interpretation: relative engagement of regulatory or cognitive control processes. A growing body of evidence suggests that increased theta power and synchronization between medial to lateral PFC occurs when the performance monitoring system detects stimuli important for learning or behavioral change (e.g., loss feedback or no-go response inhibition; Cavanagh et al., 2009; Cohen & Cavanagh, 2011; Marco-Pallares et al., 2008; Watts et al., 2018). As such, it is possible that a saturation effect may have occurred during processing loss and no-go stimuli, such that phase synchrony was universally enhanced across all subjects. As a result, there may not have been sufficient variance in the loss and no-go conditions to detect symptom change effect.

Additionally, it is necessary to address the Gain specific effects during the Gambling feedback task. One of the most common underlying traits across anxiety and depressive disorders is a tendency to experience a negativity bias (Hansen & Hansen, 1988; Ito et al., 1998). Individuals with a negativity bias are characterized by a vulnerability to psychological feedback, reflected by an increased responsiveness to aversive events, and a decreased sensitivity to anticipated or actual (Craske et al., 2023; McFarland & Klein, 2009; Noworyta et al. 2021; Pizzagalli et al., 2008) rewards. Because DLPFC is involved in the appraisal of both positive and negative information (Golkar et al., 2012; White et al., 2023), lower phase synchrony with this region during gain feedback may represent reduced regulatory ability for rewards. Therefore, by targeting and modifying negative cognitive biases toward a more positive or benign interpretation of information (i.e., CBM portion of the intervention), perhaps the current intervention is the most effective for individuals who have a negativity bias and deficit in reward-specific regulatory processes.

### Shared variance among MFT Gain and Go conditions

Finally, regarding the secondary aim of the study, results show that medial frontal theta phase synchrony during the Gain and Go conditions demonstrates shared variance in predicting treatment outcome. Although these components are elicited in different tasks, their shared variance suggests that lower synchronization between medial to lateral prefrontal regions is a reliable indicator of treatment outcomes. This is consistent with other studies in the literature that suggest MFT represents a shared process in relation to motivationally significant outcomes (Cavanagh et al., 2012; Cavanagh & Shackman, 2015; Rawls et al., 2019).

Cognitive control is supported by at least two mechanisms: proactive control and reactive control (Braver, 2012; Braver, Gray, & Burgess, 2007). Proactive control is resource intensive and reflects reward processing (e.g., gain feedback) as well as active maintenance of task goals (e.g., press the button for a Go stimulus) and is particularly suitable to tasks that elicit high cognitive demand (Braver et al., 2012). Reactive control, on the other hand, reflects the *triggering* of control mechanisms, such as retrieval of task sets or goals when conflict or adverse outcomes are identified (e.g., no-go and loss stimuli).

Directly relevant to the current findings, previous research shows the two types of control (proactive v. reactive) are differentially utilized in individuals with anxiety. Using a working memory N-back task, Fales and colleagues (2008) found that anxiety was associated with a neural signature of increased reactive control and reduced proactive control, particularly on high interference trials. Taken together with the results from the present study, it is possible that low phase synchrony in the gain and go conditions reflects less engagement of proactive control mechanisms. Since the current intervention works to reduce anxiety, a reduction in anxious symptoms may translate to increased availability of cognitive resources needed to implement proactive control. Therefore individuals with lower pretreatment engagement of *proactive* control may have more room to benefit from the current intervention.

### Amplitude

Regarding amplitude, results indicated that contrary to the hypothesis, MFT amplitude did not predict treatment response for any of the treatment groups relative to the control group. The effects were non-significant for both Theta-FN and Theta-N2 under each task condition (gains and losses, go’s and no-go’s). This difference may be substantive, such that amplitude is less sensitive than functional connectivity with lateral frontal areas. This would support the inference that it is the engagement of lateral-frontal control networks that are more predictive than the initial salience of the stimulus.

Several methodological reasons for the lack of effects for amplitude should be considered. First, it could be due to measurement methods and sample characteristics. The effects of ACC on predicting treatment outcomes, for example, may not be well detected at high temporal resolutions such as the milliseconds in the MTF activation compared to the seconds in the Blood Oxygen Level Dependent (BOLD) signal in fMRI studies. Alternatively, it could be due to a suppression effect based on the heterogeneity of the sample since the current study included individuals across a variety of diagnoses while most of the treatment response literature has been done in depressed samples. Similarly, we utilized a behavioral intervention while medication studies were more prevalent in the literature. It is possible that the direction of ACC treatment response effects may be diagnostic specific as evidenced by the few studies predicting psychotherapy outcomes (Dichter, Felder, & Smoski, 2010; McGrath et al., 2014; Hendler et al., 2003; Siegle et al., 2006, 2012). However, given the low sample sizes within each diagnostic category and treatment condition, additional tests could not be conducted to confirm or refute these potential explanations.

### Limitations and future directions

There are several limitations that should be acknowledged when interpreting the results from the current study. First, the ERN could not be assessed due to an insufficient number of errors committed in the tasks. Given its strong association with anxiety and ties to endogenous/internal sensitivities to performance monitoring, the ERN may have been particularly related to symptom improvement in the anxiety sensitivity treatment outcome measure. As such, evaluating the ERN as a predictor of treatment response represents an untapped area of research that future studies will need to examine. EEG data for testing modulations in MFT before and after the intervention was not collected in the current study design. Future research should investigate whether changes in amplitude or phase synchrony occur as a result of the intervention. If the significant predictors of treatment response were to demonstrate change after the intervention, it would provide additional evidence of treatment effectiveness as a function of targeting specific neural networks and cognitive processes for certain individuals. With regard to the non-significant amplitude results, further work could be done to assess whether this represents a lack of treatment response potential, or whether it may be the result of a suppression effect due to the heterogeneity of the sample or that amplitude treatment-related change in amplitude may be essential to assess for differences. Finally, the intervention includes multiple treatment components and a lack of behavioral data to assess the efficacy of each individual component in relation to the others. It is also unknown if the current predictors are specific to this particular type of treatment as there was a lack of a standardized treatment comparison group, such as Cognitive Behavioral Therapy or a proven pharmacological intervention. Future research should expand upon the current study by testing medial frontal theta ERPs and phase synchrony measures across multiple evidence-based interventions.

### Conclusion

Despite the above limitations, results of the current study represent a promising avenue for future research. As the first study to examine task-based medial frontal theta components as predictors of treatment response, the current findings reflect a novel contribution at the forefront of an emerging field. The observed lower baseline medial to lateral prefrontal phase synchrony during the Gain and Go conditions also provides additional support for a shared medial frontal theta process, and suggests that low engagement of regulatory and proactive control mechanisms is predictive of better response to cognitive behavioral interventions. As such, this work may ultimately lead to the improvement in treatment efficacy by serving as a target for future interventions and a method of improving individualized treatment selection.

## Data Availability

All data produced in the present study are available upon reasonable request to the authors.

